# Markerless motion capture to assess upper extremity movements in individuals with dyskinetic cerebral palsy: an accuracy and validity study

**DOI:** 10.1101/2023.08.09.23293642

**Authors:** Inti Vanmechelen, Ellen Van Wonterghem, Jean-Marie Aerts, Hans Hallez, Kaat Desloovere, Patricia Van de Walle, Annemieke I. Buizer, Elegast Monbaliu, Helga Haberfehlner

**Author notes:** **Correspondence:** Helga Haberfehlner;, Spoorwegstraat 12, B-8200 Bruges, BELGIUM.

## Abstract

**Aim:** To evaluate clinical utility of markerless motion capture (MMC) during an reaching-sideways-task in individuals with dyskinetic cerebral palsy (DCP) by determining (1) accuracy of key points tracking in individuals with DCP and typically developing (TD) peers, (2) concurrent validity by correlating MMC towards 3D-motion analysis (3DMA) and (3) construct validity by assessing differences in MMC features between a DCP and TD group.

**Method:** MMC key points were tracked from frontal videos and accuracy was assessed towards human labelling. Shoulder, elbow and wrist angles were calculated from MMC and 3DMA (as gold standard) and correlated. Additionally, execution time and variability features were calculated from key points. MMC features were compared between groups.

**Results:** Fifty-one individuals (30 DCP;21 TD; age:5-24 years) participated. An accuracy of approximately 1.5 cm was reached for key point tracking. While significant correlations were found for wrist (ρ=0.810;p<0.001) and elbow angles (ρ=0.483;p<0.001), MMC shoulder angles were not correlated (ρ=0.247;p=0.102) to 3DMA. Wrist and elbow angles, execution time and variability features all differed between groups (Effect sizes 0.35-0.81;p<0.05).

**Interpretation:** Videos of a reaching-sideways-task processed by MMC to assess upper extremity movements in DCP showed promising accuracy and validity. The method is especially valuable to assess movement variability within DCP without expensive equipment.

## Introduction

Upper extremity assessment in cerebral palsy (CP) has gained increasing interest in the past decade, with three-dimensional motion analysis (3DMA) establishing as gold standard within clinical movement assessment [1–3]. In 3DMA, markers are attached to the skin, providing accurate calculations of joint angles and spatio-temporal parameters based on biomechanical models [4]. 3DMA in CP has involved several functional tasks such as reach-grasp and drinking [4–6] or a game-like setting [7]. However, drawbacks of 3DMA are that measurements are time-consuming, they imply some discomfort for children (such as attachment and removal of skin markers), and they require expensive equipment. As a cost-effective alternative for the assessment of upper extremity movements, the use of wearable sensors has been explored within CP [8–10]. Another even more cost-effective and convenient technique could be markerless motion capture (MMC) from single camera recordings [11].

MMC is a rapidly evolving technology based on computer vision to track movement. DeepLabCut [12] is one of the recently developed open-source codes that allows users to track user-defined (body)key points in an accessible way. Within DeepLabCut, models that automatically track these key points can be trained with a small number of manually added labels. These key points are subsequently extracted as time-series of 2D pixel coordinates from the video and can be visualized as a stick figure animation. Such extracted stick figures from a single camera have previously been used as input data to develop applications in movement assessment in CP [13–16]. Examples are the early detection of CP in infants at risk based on General Movement Assessment [14,15], the prediction of gait parameters in ambulatory children with CP [13] or the automatic detection of dystonia in individuals with dyskinetic CP (DCP) [16]. However, MMC from a single camera has not yet been reported for the assessment of upper extremity movements in children with CP [11].

Current research on upper extremity assessment in CP mainly focuses on the spastic or general CP population, with underrepresentation of the DCP group. However, a fine-tuned upper extremity assessment in individuals with DCP could contribute to the indication for and evaluation of targeted treatment. For this group, assessing an upper extremity task is especially challenging due to the high variability in movement execution caused by involuntary movement and abnormal postures (i.e. dystonia and choreaoathetosis) [5,17,18]. Apart from a high movement variability, children and adults with DCP are known to perform upper extremity tasks slower, with a longer trajectory and reduced joint range of motion compared to typically developing (TD) peers [17–19]. For individuals with DCP, an easily applicable tool would be especially needed to objectively assess their movement characteristics for potential use in an natural environment. A video-based assessment - using DeepLabCut to extract key point coordinates and subsequently calculate movement features from these coordinates – could therefore be very useful in this context. A DeepLabCut model that is well-trained on upper extremity reaching tasks could be suitable to evaluate new videos without extra manual labelling.

As a first step towards a video-based assessment method of upper extremity movement within DCP, both the accuracy of key point tracking and the validity of calculated movement features need to be assessed. Therefore, the current study aims to (1) evaluate the accuracy of automated tracking of key points using DeepLabCut compared to human labels in children and adolescents with DCP and TD peers during an upper limb task, (2) validate MMC movement features towards 3DMA as gold standard (i.e., concurrent validity) and (3) assess if differences in MMC movement features between a DCP and TD group can be evaluated using MMC (i.e., construct validity).

## Method

This study is a secondary analysis of data from a multicentre study on an Instrumented Dystonia and Choreoathetosis Assessment (IDCA) protocol of upper limb movements in DCP using 3DMA, inertial sensors, common video recordings and clinical measures of dystonia and choreoathetosis [10,17,20]. Videos and 3DMA data from the IDCA study were used in the current study.

The primary study as well as the secondary use of the data was approved by the Medical Ethics Committees of the University Hospitals Leuven/KU Leuven and Vrije Universiteit Medical Center, Amsterdam. Written informed consent was obtained from participants and/or their parents.

### Participants

Participants were recruited at three movement laboratories in Belgium and one in the Netherlands: (1) Laboratory of Clinical Movement Analysis (Heder, Antwerp), (2) Clinical Motion Analysis Laboratory (UZ Leuven, Pellenberg), (3) We-Lab for Health, Technology and Movement (KU Leuven, Bruges) and (4) Clinical Motion Analysis Laboratory (Amsterdam UMC, Amsterdam). The dataset consisted of 30 participants with DCP (24 participants from the labs in Belgium and six from the Amsterdam lab). The 21 typically developing participants were all measured in the Belgian labs.

Inclusion criteria for the IDCA study for individuals with DCP were: a diagnosis of DCP, aged between 5-25 years and the ability to execute the requested tasks. Participants were excluded if they had neurological disorders other than DCP. TD participants within the same age range were recruited via personal communication. All participants from whom a frontal video recording of a reaching-sideways-task was available, were included in the current analysis. Eight repetitions from the data for each participants were extracted.

### Task execution

All participants were asked to perform a reaching-sideways-task towards a target. Start position of the hand was indicated with an elastic band on the ipsilateral knee. The reaching height was set at shoulder height (acromion). The reaching distance was determined according to arm length (from acromion to metacarpophalangeal III joint (MCP3)) using a custom-made frame to keep reaching distance constant. All tasks were performed at self-selected speed with the non-preferred or more impaired arm. A few repetitions were allowed to practice.

### Three-dimensional motion analysis (3DMA)

3DMA was performed following Upper Limb Evaluation in Motion as previously described in detail [4,17]. A brief description is provided in the supplementary material (S1).

### Markerless motion capture (MMC)

The videos were recorded frontally with a Sony video camera (HDR-CX450, Sony Europe B.V., Dublin, Ireland) fixated to a tripod at a distance of about two meters. Videos had a resolution of 1280×720 pixels and were recorded with a frequency of 25 Hz. From the target size as a known distance in the videos one pixel is estimated as 2 mm.

To avoid privacy issues federated learning was applied [21], meaning that original videodata were not pooled together during MMC, i.e. model training, evaluation and extraction of key points were performed locally and only extracted key points were aggregated for further analysis. DeepLabCut version 2.3 [12] was used for MMC, using a desktop computer with a single NVIDA GeForce RTX 3090 GPU (KU Leuven) and a Microsoft Azure’s virtual environment provided by anDREa consortium (www.andrea-cloud.eu) with a single NVIDA Tesla K80 GPU (Amsterdam UMC). Twenty frames for labelling were automatically selected using k-means. A ResNet-101-based neural network pretrained on the MPII Human pose dataset [22] provided by DeepLabCut was used to train a model for 400,000 iterations. The MPII pretrained model includes the following key points: ankle, knee, hip, shoulders, elbow, wrist, forehead and chin. MCP3 and the target were added to these predefined key points. Examples of extracted stick figures animations of a TD child and a child with DCP are available as supplementary material S2, S3. The joint angles at point of task achievement (PTA) of the second repetition were used for concurrent validity, and all eight repetitions for construct validity.

Calculation of the movement features of MMC was performed with customized code in Python (Version 3.8.15, https://www.python.org/). The code is online available together with the extracted key point data [23]. 2D frontal plane joint angles were calculated as angles between the segments hip-shoulder and shoulder-elbow (shoulder angle), between shoulder-elbow and elbow-wrist (elbow angle) and between elbow-wrist and wrist-MCP3 (wrist angle).

As measures of trajectory deviation, the overshoot of the wrist trajectory above the object (as y-distance) for each trial, as well as the area and perimeter of the convex hull [24] for all eight trials together, were calculated (Fig 1.). For all measures of trajectory deviation, x and y coordinates were normalized to the distance between start position and PTA of the wrist. For a consistent trajectory, the overshoot of the wrist was expected to be minimal, with a very small area of the convex hull and a perimeter close to two (i.e. two is the minimal distance of the trajectory from start to PTA and back).

**Fig. 1.**
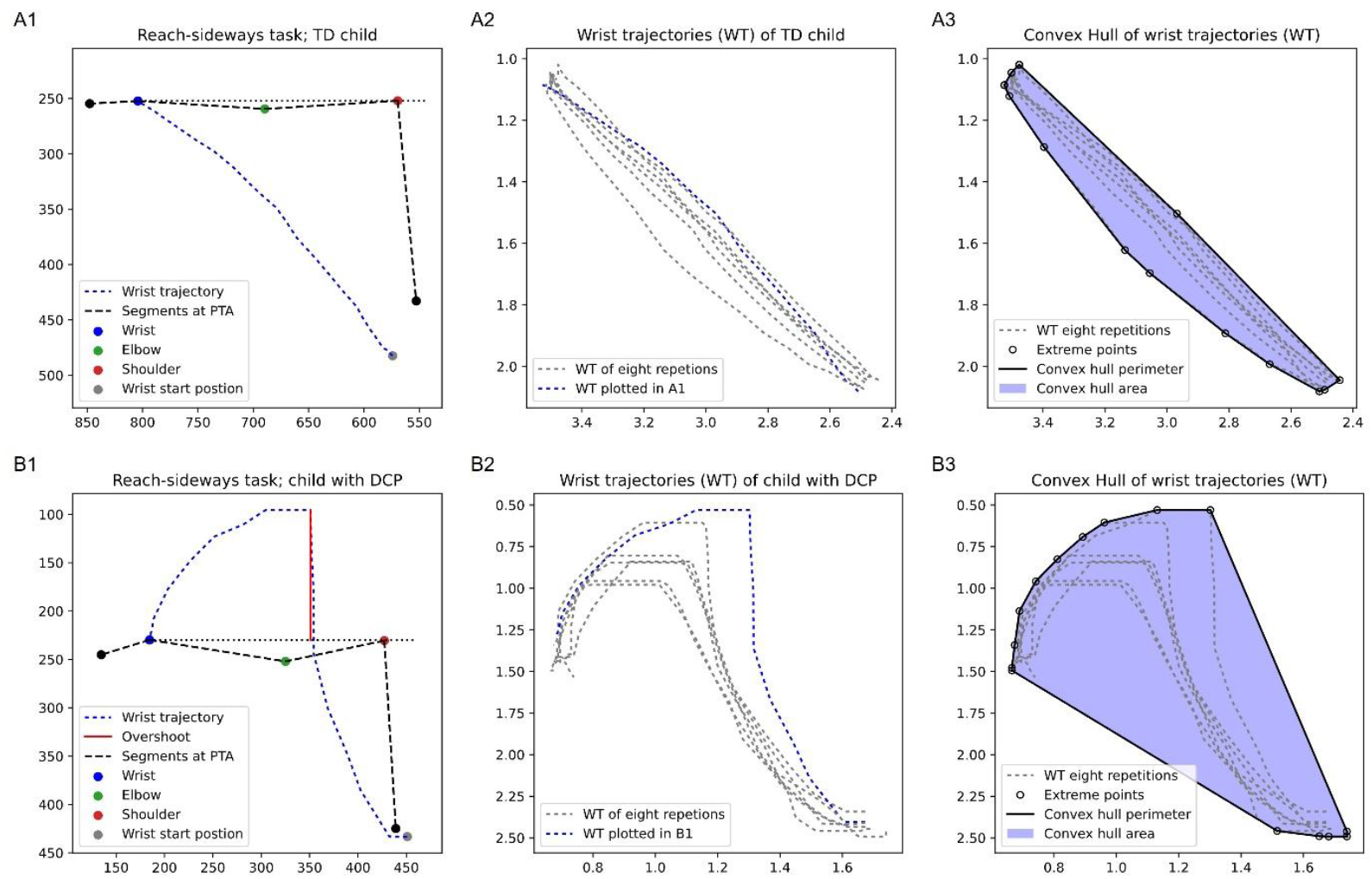
Visualization of feature extraction for a reaching-sideways-task using extracted key points from a frontal video by DeepLabCut. An example from a typically developing child (TD) (A) and a child with dyskinetic cerebral palsy (DCP) (B) are presented. In A1 and B1 the wrist trajectory (blue) of one repetition, the arm position at the point of task achievement (PTA), as well as the overshoot (red) is plotted. Note that in the example of the TD child (A1) no overshoot was observed. In A2, B2: The wrist trajectory of eight performed repetitions are plotted, within blue the repetition from A1, B1 respectively. A3, B3 shows the perimeter and area from the convex hull of eight repetitions. The area and perimeter of the convex hull were calculated as an indication of movement variability. The stick figure videos of these two examples are presented in the supplementary material (S2, S3).

### Split dataset to assess tracking accuracy

To assess tracking accuracy, the dataset was split in a training (70%), validation (5%) and two test datasets (test-dataset-I (15%), test-dataset-II (10%)). The test datasets were kept separate during training of the model. Twenty participants with DCP and 17 TD peers from Belgium (measured in three different labs) were included in the training and validation dataset for model development. The test-dataset-I consisted of four individuals with DCP (randomly selected taking into account an equal spread of MACS levels) and four TD individuals from the Belgium data. The test-dataset-II consists of all six participants from Amsterdam.

Tracking accuracy was calculated by the mean absolute error (MAE) (i.e., coordinates of manually labelled key points *versus* coordinates of key points predicted by the model in pixels). MAEs were calculated with a p-cutoff of 0.8 (i.e., leaving out predictions with a low likelihood). This low likelihood was mainly due to occlusion of body parts or motion blur. A MAE of about 7.5 pixels (approximately 1.5 cm) was considered sufficiently accurate for further analysis. In addition, overlay videos of original videos with tracked key points were visually inspected to assess tracking accuracy and possible outliers. As tracking accuracy turned out to be lower in the test datasets compared to the development dataset (training and validation dataset), a maximum of five outlier frames within each video were extracted in DeepLabCut. These outliers were subsequently relabelled and the model was retrained, prior to further data analysis.

### Statistical analysis

To establish concurrent validity, the MMC movement features and 3DMA movement features were individually correlated. For construct validity of MMC, hypothesis testing was used assessing known-group differences [25]. Following differences were expected to be found in individuals with DCP compared to TD peers: (1) a longer execution time (extracted from movement cycles), (2) reduced joint angles and (3) higher variability (measured as standard deviation between trials for the joint angles and trajectory deviation by mean overshoot and convex hull).

For all variables, normality was tested using the Shapiro–Wilk test. As normality was not confirmed, correlations were calculated with Spearman correlation coefficients (ρ) on the pooled data of the DCP and TD group. Correlations ranging from 0.00-0.25 were interpreted as low or no relationship, 0.25-0.50 as fair, 0.50-0.75 as moderate to good and above 0.75 as excellent [26]. Between-group comparisons was performed using Mann Whitney U-Tests. Effect size (ES) was calculated by 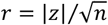. Statistics were calculated with SPSS 22.0 (IBM Corp., Armonk, NY, USA). A p-value of <0.05 was considered statistically significant.

## Results

### Participants

Characteristics of the 30 participants with DCP were: age 17.3 ± 5.3 (mean ± standard deviation) years; 10 females/20 males; Manual Ability Classification System (MACS): I (n = 3), II (n = 13), III (n = 12), IV (n = 2); and of the 21 TDs: age 17.3 ± 4.2 years; 16 females/5 males. Within the DCP group, 18 participants performed the task with the left arm, 12 with the right arm. Within the TD group, 18 participants performed the task with the left arm and three with the right arm.

### Tracking accuracy

The MAE within the unseen data (both test datasets) was overall twice as high compared to the seen data (training dataset: 2.66 pixels, validation dataset: 2.91, test-dataset-I: 6.44 pixels and test-dataset-II:7.73 pixels) (Table 1). Notwithstanding, most key points revealed an accuracy close to the predefined 7.5 pixels, with the exception of the hip within the test-dataset-I (MAE=14.13 pixels) and the MCP3 test-dataset-II (MAE=10.59 pixels). Visual inspection of the overlay videos revealed problems with the tracking accuracy in one video in the test-dataset-I concerning tracking of the hip due to obesity. For two videos in the test-data-set-II, tracking accuracy was lower due to an assisting person in view in the videos. Interestingly, skin tone differences of participants in the dataset did not seem to effect tracking accuracy (as visually assessed between participants with pale and dark skin tones).

**Table 1:** Average of MAE in pixels between manual human label and label predicted by DeepLabCut of the executing body half with an applied p-cutoff of 0.8.

| Key points | Development of model |  | Evaluation of model |  |
| --- | --- | --- | --- | --- |
|  | Training dataset | Validation dataset | Test-dataset-I | Test-dataset-II |
| *Hip* | 3.10 | 3.16 | 14.13 | 3.26 |
| *Shoulder* | 2.56 | 3.12 | 8.76 | 7.40 |
| *Elbow* | 2.90 | 2.61 | 7.76 | 5.18 |
| *Wrist* | 2.59 | 2.80 | 7.22 | 6.84 |
| *MCP3* | 2.51 | 3.81 | 6.24 | 10.59 |
| *Target* | 2.40 | 2.31 | 2.98 | 8.50 |
| *Total* | 2.66 | 2.91 | 6.44 | 7.73 |

MCP3 = metacarpophalangeal III joint; MAE = Mean absolute error

### Concurrent validity

For five of the participants with DCP, no 3DMA data was available (technical reasons in three cases and problems with marker visibility due to impairment in two cases). This cases were not included in the calculation for concurrent validity. 3DMA shoulder elevation showed a low, not significant correlation to MMC shoulder angle (ρ=0.247 [CI: -0.059-0.511]; p=0.102), while elbow flexion/extension and wrist flexion/extension from 3DMA were significantly correlated to elbow and wrist angle from MMC, respectively (ρ=0.483 [CI: 0.215-0.683]; p<0.001) and ρ=0.810 [CI: 0.672-0.893]; p<0.001) (Fig 2.)

**Fig 2.**
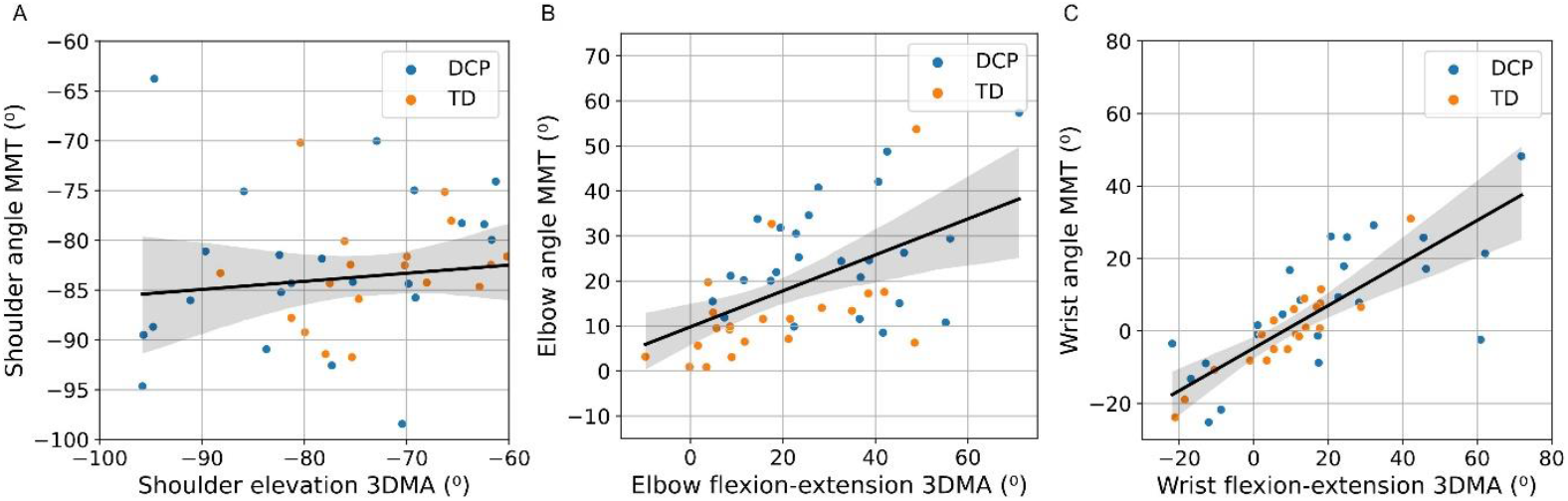
Correlation between shoulder elevation (A), elbow flexion-extension (B) and wrist-flexion extension (C) measured by 3D-motion analysis (3DMA) and shoulder, elbow and wrist angle measured by markerless motion capture (MMC) from 2D videos.

### Construct validity

Median and inter-quartile ranges of MMC features are presented in Fig 3. Significant differences between TD and DCP groups were found for the wrist flexion/extension angles (ES=0.35;p=0.016) and elbow flexion/extension angles (ES=0.50;p<0.001), with more flexion in the wrist and elbow in the DCP group at PTA (Fig 3.C,E). Shoulder angles at PTA did not differ between groups (ES=0.04;p=0.797) (Fig.3.A). Participants with DCP performed the task on average slower in comparison with TD peers (ES=0.56;p<0.001) (Fig.3.G). All features of movement variability significantly differed between groups, with higher values (i.e., more variability) in the DCP group: Standard deviation of shoulder angle (ES=0.53), elbow angles (ES=0.49) and wrist angles (ES=0.57) at PTA of eight trials (all p<0.001) (Fig 3.B,D,F), overshoot (ES=0.75), convex hull area (ES=0.81) and the convex hull perimeter (ES=0.76) (all p<0.001) (Fig. 3.H.I,J).

**Fig 3.**
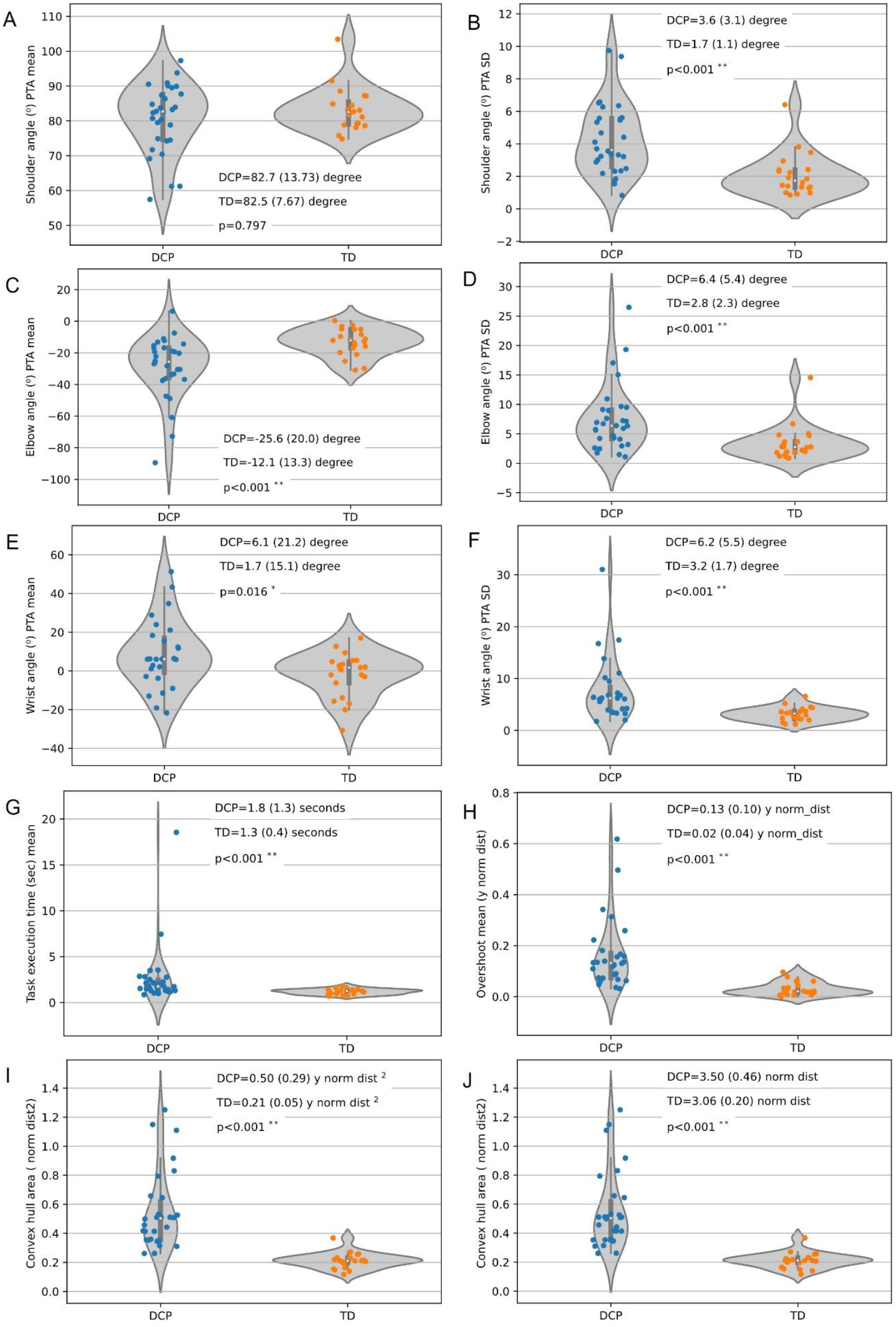
Differences between individuals with dyskinetic cerebral palsy (DCP) and typically developing peers (TD) for mean joint angles at point of task achievement (PTA) and the standard deviations (SD) from eight repetitions (A-F), mean execution times (G) and mean overshoot (H), convex hull area (I) and convex hull perimeter (J). Individual values as well as medians and inter-quartile ranges are presented. Within the violin plot median is plotted as white spot, the inter-quartile range as black bar and the upper and lower adjacent value as thin line.

## Discussion

The clinical utility of a video-based assessment using MMC during a reaching-sideways-task for upper extremity movements in individuals with DCP was tested within the current research. Within this approach DeepLabCut was used to extract key point coordinates from videos and subsequently a selected set of movement features was calculated from these key points. The calculated features represent clinically relevant movement features in DCP, such as joint angles, execution time and movement variability. Overall accuracy and validity of the proposed method was promising, opening possibilities to facilitate remote movement analysis in DCP.

A similar tracking accuracy of key points in the training and validation dataset, showed that within a seen dataset accuracy is adequate and key points can be used for subsequent calculation of movement features. However, within unsee/new participants and/or environments, the model did not yet accurately track body parts of all subjects. In the current study, this aspect was solved by adding extra manual labels for a few participants and retrain the model. The current workflow - using federated learning which requires no pooling of videodata at one location - can facilitate multicenter collaborations for the relatively small population of individuals with DCP. Still, it is expected that a generalizable DeepLabCut model for a reaching-sideways-task will be reached in the near future with a larger and more variable video dataset (e.g. various background, clothing and body forms). Within similar research, a generalizable DeepLabCut model for tracking movements in infants was achieved with a training dataset of 100 participants [15]. In addition, ongoing developments will accelerate accurate and fully-automated extraction of key points from a single camera and key points may even include 3D information from smartphones-cameras with depth sensors in the future [11].

Concurrent validity was sufficient for wrist angles at PTA measured by MMC, whereas correlations between 3DMA and MMC were fair for the elbow angles and low for the shoulder angles. Most likely the lack of 3D representation within the MMC angle features have affected correlations at PTA due to out-of-plane movements, such as combined shoulder rotation and elevation.

Nevertheless, the expected between-group differences concerning joint angles necessary to establish construct validity of MMC angles at PTA were found, meaning impaired elbow and wrist extension in the DCP group compared to TD. For the MMC shoulder angle, no significant differences were found, which corroborates previous research failing to find deviant shoulder elevation angles during a reaching-sideways-task [17], although these differences were found within tasks in the vertical plane such as simulating drinking and reach-and-grasp vertically [17,19]. Construct validity was confirmed for task execution time and all movement variability features measured by MMC. The newly introduced features - convex hull perimeter, convex hull area and overshoot indicating trajectory deviation - all revealed an ES above 0.75 for the group difference between DCP and TD group.

As high variability in movement execution caused by involuntary movement and abnormal postures hampers activities of daily living, rehabilitation and pharmaceutical treatment often target to reduce these involuntary movements [27]. Personalized treatments would benefit from frequent assessments with the possibility to extract objective movement features. MMC is highly promising to be developed towards an easily applicable and sensitive tool to assess upper extremity movements in DCP in clinical practice. For future application, this approach could be embedded in a smart-phone application for use as an out-of-clinics tool within outpatients clinics for rehabilitation, physiotherapy and occupational therapy or within a home-based setting. The reaching-sideways task, used in the current study can be extended with other daily activity upper extremity tasks.

To clinically use MMC to assess upper extremity movements in individuals with DCP, a reference data set of TD peers is necessary taking into account possible effects of age. With such a data set, a cut-off value for the distinct MMC features per age group can be defined. This would then allow the assessment of pre-post differences as distance to this cut-off value. Furthermore, psychometric properties of current MMC features need to be further evaluated including responsiveness, reliability and validity towards clinical scales (e.g. the Dyskinesia Impairment Scale [28]), clinical arm-hand function tests (e.g. Jebsen-Taylor Test [29]) and questionnaires (e.g. ABILHAND-Kids [30]).

## Conclusion

This study was the first step towards a video-based method to assess upper extremity movements in children and adolescents with DCP. Here, accuracy and validity of such an approach is presented. MMC – especially at the time that fully-automated key point tracking is reached – offers possibilities for frequent evaluation of upper extremity function in a group of patients showing high movement variability.

## Supporting information

S1 Brief description of three-dimensional motion analysis (3DMA

S3 Video: Example of extracted stick figure of a child with dyskinetic cerebral palsy (DCP) during a reaching-sideways task

## Supporting information

**S1** Brief description of three-dimensional motion analysis (3DMA)

**S2** Video: Example of extracted stick figure of typically developing (TD) child during a reaching-sideways task

**S3** Video: Example of extracted stick figure of a child with dyskinetic cerebral palsy (DCP) during a reaching-sideways task

## Data Availability Statement

The data and code used in the current study is available online https://doi.org/10.48804/BZM9SJ

## Acknowledgement

The authors would like to thank the participants and their parents for their time and effort in participating in the data collection. The authors acknowledge the contribution to the presented work of Thomas Swinnen, who contributed as master student physiotherapy to the study.

