## Supplementary material for "Markerless motion capture to assess upper extremity movements in individuals with dyskinetic cerebral palsy: an accuracy and validity study": S1 Brief description of three-dimensional motion analysis (3DMA

S1 Supplementary material

*Brief description of three-dimensional motion analysis (3DMA)*

For a detailed description of 3DMA see Jaspers et al. and Vanmechelen et al. [1,2]. The related software matlabcode is available online https://github.com/u0078867/ulema-ul-analyzer.

In brief: Seventeen reflective markers were placed over the body in five clusters; two cuffs of four markers were placed on the upper arm and forearm, one cluster of three markers was placed on the hand and two tripods with three markers were placed on the trunk and the scapula. Five segments were thus included (trunk, scapula, humerus, forearm and hand) resulting in four joints considered for 3DMA (scapulothoracic (scapula), humerothoracic (shoulder), elbow and wrist).

Movement cycles of 3DMA were visually identified and segmented in Vicon Nexus (Vicon Motion Systems, Oxford Metrics, UK). One movement cycle was defined from hand on ipsilateral knee to point of task achievement (PTA), where PTA was considered the final point of the reach cycle.

Calculation of joint angles and spatiotemporal parameters for 3DMA were calculated with Upper Limb Evaluation in Motion Analysis (U.L.E.M.A) protocol using Matlab (MATLAB R2019B, The MathWorks Inc., Natick, Massachusetts) [1,2].

[1] Vanmechelen I, Bekteshi S, Konings M, Feys H, Desloovere K, Aerts JM, et al. Psychometric properties of upper limb kinematics during functional tasks in children and adolescents with dyskinetic cerebral palsy. PLoS One 2022;17. https://doi.org/10.1371/JOURNAL.PONE.0266294.

[2] Jaspers E, Feys H, Bruyninckx H, Harlaar J, Molenaers G, Desloovere K. Upper limb kinematics: Development and reliability of a clinical protocol for children. Gait Posture 2011;33:279–85. https://doi.org/10.1016/J.GAITPOST.2010.11.021.
